# Substance Use Disorder Risk Assessment: Preventing Use Disorder

**DOI:** 10.1101/2022.12.08.22283157

**Authors:** Karen E. Arscott, Donna M. Eget, Maria C. Marcos, Brian J. Piper

## Abstract

**Importance:** Current screening tools for Alcohol Use Disorder (AUD) or Substance Use Disorder (SUD) discover this disease at a late stage.

**Objective:** The goal of this study was to develop a simple prevention screening tool for patients who may be prone to develop AUD and/or SUD prior to the development of addiction.

**Design:** The study involved a self-administered survey type study using a cross-sectional design and was carried out in the spring of 2020 over a one-month period.

**Setting:** This study was completed at an urgent care facility which supports a separate Medication Assisted Treatment (MAT) clinic. Those patients who presented to the MAT clinic (SUD group) were seen in a separate area that the patients presenting for urgent care (Control group).

**Participants:** Participants (N = 259) were voluntarily recruited from MAT and Urgent care patients: Patients receiving acute care were assigned to the Control group (N = 126, 50.8% female, 5.7% non-white, 27.2 age < 34)) and those receiving treatment for SUD were assigned to the MAT group (N =133, 40.8% female, 4.8% non-white, 36.8% <34)).

**Main Outcome and Measure:** The survey questioned demographics (4 items), risk factors for AUD/SUD (6 items), information about first alcohol/opioid experiences (16 items), and factors for seeking AUD/SUD treatment and recovery (2 items). Feelings were categorized as positive (e.g., euphoria, happiness, self-confident), neutral (e.g., nothing, normal), or negative (e.g., depressed, sad, sick).

**Results:** The MAT group felt more positive feelings with first usage of alcohol and opioids compared to the control group (p<.001). With first usage of opioids specifically, MAT (0.13 + 0.04) and Control (0.29 + 0.07, p <.001) groups differed. Over half (55.3%), of the MAT participants reported feeling self-confident with first use of alcohol while only 29.7% of the Control reported this (p<.001). Over three-fifths (63.7%)of the MAT group reported feeling of euphoria with the first usage of opioids compared to one-tenth (9.8%) in the Control group (p<.001).

**Conclusions:** This retrospective cross-sectional report shows the first affective responses to substances may predict risk for future SUD and could be used as a prevention screening tool. Reporting positive feelings with first usage of alcohol and opioids could be used as a screening tool for patients who may be more prone to developing AUD and SUD.

**Key Points:** *Question:* Is there a way to identify a person at risk for developing a substance use disorder?

*Findings:* In this retrospective cross-sectional study involving 259 participants it was significantly demonstrated that the first affective responses to recreational drugs may predict risk for future drug misuse potentially leading to SUD. The MAT group felt more positive feelings with first usage of alcohol and opioids compared to the comparison group (p < .001).

*Meaning:* If a patient develops a euphoric response to initial experience with a substance, they are significantly more likely to develop a substance use disorder.

## INTRODUCTION

The current screening tools used to determine if a person has an Alcohol Use Disorder (AUD) or Substance Use Disorder (SUD) are unfortunately designed to discover these diseases at a late stage. As with most diseases the best method to avoid AUD or SUD would be prevention. To date, there is no research describing a tool such as this study proposes. A study conducted in 2016 examined the question of whether there is a difference between prescribed opioids or those taken experimentally and provided a starting point for investigating the feelings persons feel the first time an opioid is used^1^. This “initial feeling” is the basis for our research project.

Tools currently in use for AUD are repurposed for other substances. These tools: CAGE (Acronym Questionnaire)^2^ ; Michigan Alcohol Screening Test (MAST)^3,4^ ; Alcohol Use Disorder Identification Test (AUDIT)^5^ ; Current Opioid Misuse Measure (COMM)^6^ ; Rapid Opioid Dependence Screen (RODS)^7^ ; Leeds Dependence Questionnaire (LDQ)^8^ ; Tobacco, Alcohol, Prescription medications and other Substance (TAPS) Tool^9^ ; Opioid Risk Tool (ORT)^10^ ; and Alcohol Screening and Brief Intervention (ASBI)^11^ are all designed to identify an AUD/SUD after it is an addiction and a serious problem. The purpose of this study was to uncover an earlier point of identification for prevention – perhaps as a “Preaddiction” flag as discussed elsewhere^12^.

While working at a Medical Assisted Treatment (MAT) clinic, many patients with AUD/SUD described their initial experience with alcohol or opioids as “great”, “best I have ever felt”, “finally felt normal”, “amazing”, etc. These descriptions are different than many other persons who have described their initial experience as “nauseating, terrible, or no help with pain.”

The hypothesis being tested is that persons with potential for AUD and/or SUD have a predisposition that can be determined simply by asking the question how they felt with their initial substance experience.

## METHODS

### Ethical Oversight

This study was approved by the Geisinger Health System Institutional Review Board.

### Study Setting and Patients

This study involved a self-administered survey using a cross-sectional design. The researchers used a simple group of questions administered to both the SUD group (those with known SUD) and the Control group (persons presenting to the clinic for treatment other than SUD). Those patients that present to the MAT Clinic (SUD group) were seen in a separate area than the patients coming for urgent care (Control group), and therefore the respective surveys were presented at the two different areas within the clinic. Potential study participants were identified at a local MAT clinic and urgent care facility during their standard care visits in either the MAT clinic or as persons requiring another type of medical care at the urgent care.

Participants (N = 259) were recruited from an urgent care clinic and received either acute care or medical treatment for SUD. Patients receiving acute care were assigned to the comparison group (N = 126, 50.8% female, 5.7% non-white, 27.2% age < 34) and those receiving treatment for SUD were assigned to the Medical Assisted Treatment (MAT) group (N =133, 40.8% female, 4.8% non-white, 36.8% age < 34).

The introductory description includes an Information Sheet describing the study and containing all elements of an informed consent form. The data collected was non-identifiable. The Information Sheet also included a return phone number to call should the subject have any questions or wish to withdraw from the study. Each participant was provided a unique study number for the subject to reference when requesting their study data be removed from the study.

The study began May 1, 2021and the last participant was enrolled May 31, 2021..

### Outcomes

The introductory description includes an Information Sheet describing the study and containing all elements of an informed consent form. The data collected was non-identifiable. The Information Sheet also included a return phone number to call should the subject have any questions or wish to withdraw from the study. Each participant was provided a unique study number for the subject to reference when requesting their study data be removed from the study.

Surveys included questions about demographics (four items), risk factors for AUD/SUD (six items), information about their first alcohol and opioid experiences (sixteen items), and factors for seeking AUD/SUD treatment and recovery (two items). There were items about first-time usage and participants were asked to select the emotions that they experienced. Feelings were categorized as positive (e.g., euphoria, happiness, self-confident), neutral (e.g., nothing, normal), or negative (e.g., depressed, sad, sick).

### Sample Size

259 participants were recruited from a local urgent care clinic in which people receive acute care and medical treatment for SUD. Of the patients receiving acute care at the clinic, 126 participants (50.8% identified as female, 36.8% age ≤ 34, and 5.7% identified as a non-white race) were placed into the comparison group. Of the clinic patients receiving treatment for SUD, 133 participants (40.8% identified as female, 27.2% age ≤ 34, and 4.8% identified as a non-white race) were placed into the Medical Assisted Treatment (MAT) group.

### Statistical Methods

Responses were collected and entered into *Systat*, version 13.1 for analysis. Comparisons between the MAT and Control groups were made with t-test for parametric variables and chi-square or Fisher’s exact test (N < 5) for non-parametric variables.

## RESULTS

### Participant Demographics and Personal Medical History

Over half (52.4%) of the 133 person MAT group had less than or the equivalent than a high school education, significantly higher than the comparison group (31.6%, p < .001). Almost a third (32.3%) of the MAT group reported that they were not content with their current situation (p <.001). Participants in the MAT group reported incidence of family history of AUD, illegal SUD, and prescription SUD that was significantly higher than the comparison group. The MAT group reported significantly increase personal history of AUD, illegal SUD, and prescription SUD compared to the comparison group (p < .001). A third (32.5%) of the MAT participants stated that they had a forced sexual experience in childhood (p < .001). One-fifth (19.8%) of the comparison group and two-ninths (22.3%) of the MAT group reported a personal history of ADD, OCD, bipolar, and schizophrenia. Four-ninths (43.5%) of the MAT group and one-third (32.8%) of the group had a history of depression.

The MAT group felt more positive feelings with first usage of alcohol and opioids compared to the comparison group (p < .001). With first usage of opioids specifically, MAT (0.13 + 0.04) and Comparison (0.29 + 0.07, p < .001) groups differed. Over half (55.3%), of the MAT participants reported feeling self-confident with first use of alcohol while only 29.7% of the Comparison reported this (p < .001). Over three-fifths (63.7%) of the MAT group reported feeling euphoria with the first usage of opioids compared to one-tenth (9.8%) in the Comparison group (p < .001).

### Alcohol Usage and Feelings with First Exposure

In evaluating the MAT and comparison group, 14.5% of the MAT group had taken alcohol younger than the age of 10 compared to 0.8% of the comparison group (p < .001). There was not a significant difference in alcohol type and source between both groups.

The MAT group overall felt more positive feelings with first usage of alcohol compared to the comparison group (p < .001). Euphoria was experienced in four-tenths (39.0%) of the MAT group and in one-eighth (12.5%) of the comparison pain (p < .001). Over half (55.3%) of the MAT participants reported feeling self-confident with first use of alcohol while only a little over one-quarter (29.7%) of the comparison group reported this feeling (p < .001). Feelings of happiness and well-being were significantly higher (p < .05) in the MAT group than in the comparison group. Feelings of relief, being accepted, strong, loveable, good enough, and focused were not experienced in either group with first time use.

There was not a significant difference in reported negative feelings with first time use (p = .078) in both groups.

Participants in the comparison group (0.61 + 0.06) encountered more neutral feelings associated with first alcohol use than the MAT group (0.42 + 0.05, p < .05). About three-tenths (31.3%) of the comparison group endorsed feeling normal, while only one-fifth (19.5%) of the MAT group reported this feeling (p < .05). There was a negligible difference between the MAT group (29.7%) and comparison group (22.8%) when reporting feeling nothing.

### Opioid Usage and Feelings with First Exposure

The age of first opioid use was not significant between the comparison group and the MAT group. In the comparison group, all the participants reported taking a non-heroin opioid during their first use. Specifically with first opioid use, over half (53.7%) of the MAT group were given their first opioid from a non-provider (i.e., friend, family member) and less than one-sixth (15.9%) of the comparison group received their first opioid from a non-provider (p < .001).

The MAT group felt more positive feelings with first opioid use compared to the comparison group (p < .001). Except for feeling good enough, all the positive classified feelings were significantly increased in the MAT group (Table 3). For example, almost two-thirds (63.7%) of the MAT group reported feeling euphoria with the first usage of opioids compared to one-tenth (9.8%) in the Comparison group (p < .001).

**Table 1:**
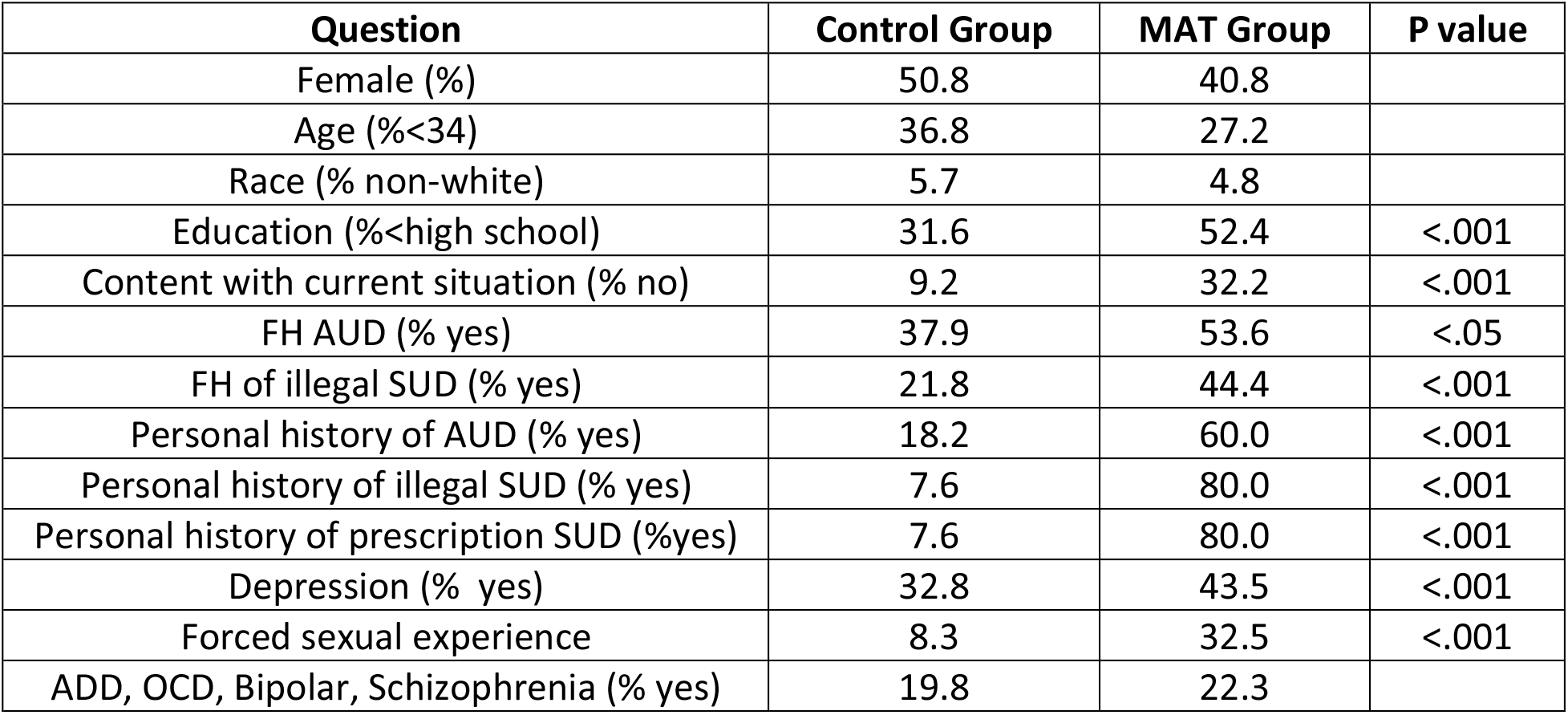
Demographics and History of Medication Assisted Treatment (MAT N=133) and Control (N=126) groups

**Table 2:** Alcohol Use and Feelings in the Medication Assisted Treatment (MAT, N=133) and control (N=126) groups

| Question | Control Group | MAT Group | P Value |
| --- | --- | --- | --- |
| Age at first alcohol <10 (%yes) | 0.8 | 14.5 | <.001 |
| Age of First Alcohol <15 (%yes) | 30.8 | 60.5 | <.001 |
| Alcohol Type -beer (%) | 67.7 | 72.6 |  |
| Alcohol type – liquor (%) | 67.7 | 72.6 |  |
| Alcohol Source – friend (%) | 49.6 | 42.7 |  |
| Alcohol Source – Self (%) | 34.9 | 44.4 |  |
| Alcohol Source – parent (%) | 7.0 | 4.0 |  |
| **Feelings** |  |  |  |
| Positive (mean + SD) | 0.95 (1.02) | 1.63 (1.35) | <.001 |
| Euphoria (%) | 12.5 | 39.0 | <.001 |
| Happiness (%) | 40.6 | 55.3 | <.05 |
| Self-Confident (%) | 29.7 | 47.2 | <.001 |
| Well-Being (%) | 11.7 | 21.2 | <.05 |
| Negative (mean + SD) | 0.27 (0.57) | 0.42 (0.76) | .078 |
| Weak (%) | 0 | 0 |  |
| Sick (%) | 21.1 | 26.8 |  |
| Depressed (%) | 1.6 | 7.3 | .055 |
| Sad (%) | 3.9 | 7.3 |  |
| Neutral (mean + SD) | 0.61 (0.63) | 0.42 (0.60) | <.05 |
| Normal (%) | 31.2 | 19.5 | <.05 |
| Nothing (%) | 22.8 | 29.7 |  |

**Table 3:** Opioid Use and Feelings in the Medication Assisted Treatment (MAT, N=133) and control (N=126) groups

| Question | Control Group | MAT Group | P Value |
| --- | --- | --- | --- |
| Age of first opioid use <15 (%) | 4.7 | 21.6 |  |
| Age of first opioid use >15 (%) | 95.3 | 78.4 |  |
| Type: Heroin (%) | 0 | 15.8 | <.001 |
| Source: Provider (%) | 84.1 | 46.3 | <.001 |
| **Feelings** |  |  |  |
| Positive (mean + SD) | 1.3(1.06) | 3.04 (0.2) | <.001 |
| Euphoria (%) | 9.8 | 63.7 | <.001 |
| Happiness (%) | 8.3 | 60.3 | <.001 |
| Self-Confident (%) | 3.8 | 41.3 | <.001 |
| Well-Being (%) | 3.0 | 23.0 | <.001 |
| Relief (%) | 23.3 | 42.9 | <.001 |
| Accepted (%) | 0 | 13.5 | <.001 |
| Strong (%) | 0.8 | 20.6 | <.001 |
| Loveable (%) | 1.5 | 7.9 | <.05 |
| Good enough (%) | 6.0 | 7.1 |  |
| Focused (%) | 2.3 | 19.8 | <.001 |
| Negative (mean + SD) | 0.29 (0.07) | 0.13 (0.42) | <.001 |
| Weak (%) | 3.0 | 0.8 |  |
| Sick (%) | 9.8 | 8.7 |  |
| Depressed (%) | 0.0 | 1.6 |  |
| Sad (%) | 0.8 | 1.6 |  |
| Neutral (mean + SD) | 0.06 (0.56) | 0.53 (0.72) |  |
| Normal (%) | 8.3 | 19.8 | <.05 |
| Numb (%) | 11.3 | 28.6 | <.001 |
| Nothing (%) | 8.3 | 4.0 |  |

**Table 4:** Alcohol and Opioid Feelings Summative in the Medication Assisted Treatment (MAT, N=133) and control (N=126) groups

| Feelings | Alcohol | Opioid | P Value |
| --- | --- | --- | --- |
| MAT Positive (Mean + SD) | 1.64 (2.05) | 3.09 (2.05) | <.001 |
| MAT Negative (Mean + SD) | 0.42 (0.83) | 0.12 (0.83) | <.001 |
| MAT Neutral (Mean + SD) | 0.41 (0.82) | 0.54 (.82) | 0.08 |
| Control Positive (mean +SD) | 1.02 (1.31) | 1.26 (1.31) |  |
| Control Negative (mean _ SD) | 0.34 (0.64) | 0.30 (0.64) |  |
| Control Neutral (Mean + SD) | 0.48 (0.81) | 0.61 (0.81) |  |

Furthermore, there was a significant difference in overall negative feelings with first opioid usage MAT participants (0.13 + 0.04) and comparison participants (0.29 + 0.07, p < .001). However, there was not a significant difference shown with comparing individual feelings that were classified as negative.

There was not a significant difference in reported neutral feelings with first time use in both groups. Over a quarter (28.6%) of the MAT group felt numb with their first opioid use, while just over a tenth (11.3%) of the comparison group reported this feeling (p < .001). A fifth (19.8%) reported feeling normal in the MAT group and a twelfth (8.3%) of the comparison group described feeling normal with first opioid use (p < .05). There was an insignificant difference between the MAT group (4.0%) and comparison group (8.3%) when reporting feeling nothing.

## DISCUSSION

This novel retrospective cross-sectional study shows that the first affective responses to recreational drugs may predict risk for future drug misuse potentially leading to SUD. Reporting positive feelings with first usage of alcohol and/or opioids could be used as a screening tool for patients who may be more prone to developing AUD and/or SUD. Group differences were generally less pronounced for neutral or negative feelings. This is an important methodological development which overcomes limitations with past instruments.^2-11^

There were over one-hundred deaths in 2021 in the US from drug overdoses.^13^ Similarly, there were over 140,000 deaths per year in the US due to excessive alcohol use.^14^ Although there are many FDA approved pharmacotherapies for AUD and opioid use disorder (but not stimulant use disorder), addiction is a relapsing and remitting disease. There are substantial individual differences in treatment response. For example, the number needed to prevent a return to drinking was twelve for acamprosate and twelve for naltrexone.^15^ Prevention of AUD and SUD, perhaps using instruments like that described in this report, should be a pressing public health priority.

## Data Availability

All data produced in the present study are available upon reasonable request to the authors

## Limitations

Although our sample size (N = 259) was sufficient to detect statistical significance across multiple areas, an increased number of participants may have strengthened this report. In addition, we made the decision to consider “normal” as a neutral feeling. If we had worded the question differently, this may have fallen on the positive side. If a participant felt “abnormal” and the use of a substance allowed the feeling of “normalcy” it would be positive. Future research will be necessary to further refine this instrument including with a more diverse sample (e.g., non-English speakers)

## Conclusions and Relevance

The primary goal of this retrospective cross-sectional study is to discover a simple screening tool for AUD/SUD prevention. The defined hypothesis was supported by the data collected and analyzed. With mortality from the opioid crisis escalating despite MAT^13^ and a wide selection of screening tools^2-11^ it is evident that prevention of the disease is required to change the trajectory of morbidity and mortality. We are cautiously optimistic that novel instruments to uncover a “preaddiction state”^12^ could contribute to precision medicine. Whenever a patient is prescribed an opioid for any reason and at any age the initial “feeling” that patient experiences should be noted. If their initial general mood is one of euphoria or contentment that individual should be cautioned about the potential for developing a substance use disorder. It could be documented as a positive “preaddiction” screen and so careful prescribing of addicting medications would be warranted. This screening tool could be widely shared in media, schools, medical/surgical offices, dental offices, and pediatric offices.

## Acknowledgements

The authors are grateful to the participants of this study who agreed to take time to respond to the survey questions and share their humanity. Also grateful to Geisinger Commonwealth School of Medicine and The Behavioral Health Institute for covering costs of research assistant and the statistical analysis. Also, wish to acknowledge and thank The Institute for reviewing the responses and analyzing the data.

## Notes

### Competing Interest Statement

The authors have declared no competing interest.

### Funding Statement

This study was funded by The Behavioral Health Institute at Geisinger Commonwealth School of Medicine

### Author Declarations

Members of Institutional Review Board (IRB) of Geisinger Health Systems reviewed and approved your research protocol under [45 CFR 46.110(b)(1) expedited review Category 7: Research on individual or group characteristics or behavior (including, but not limited to, research on perception, cognition, motivation, identity, language, communication, cultural beliefs or practices, and social behavior) or research employing survey, interview, oral history, focus group, program evaluation, human factors evaluation, or quality assurance methodologies on 04/19/2021 Please note the following information about your IRB approval: Approval Date: 04/19/2021 Risk Assigned: Minimal Risk Approved Subject Screening: 500 Approved Subject Enrollment: 240 Approved PHI Elements: No Identifiers collected Consent/Authorization Process: Waiver approved under 45 CFR 46.117 (c) 1 or 2/ 21 CFR 56.109 (c)1 HIPAA Authorization for research approved under 45 CFR 164.508 (a) (1) Sponsor: Clinic Research Fund - Research Assistance & Support, GCSOM Behavioral Health Initiative

